# The Impact of Fatty Acid Reporting Methods on Associations with Cardiometabolic Biomarkers

**DOI:** 10.1101/2025.11.19.25340593

**Authors:** Brian Hallmark, Manja M. Zec, Laurel Johnstone, Carrie S. Standage-Beier, Susan Sergeant, Justin Snider, J. Thomas Brenna, Floyd H. Chilton

## Abstract

**Background:** Global dietary guidelines for polyunsaturated fatty acids (PUFAs), especially linoleic acid (LA) and its metabolite arachidonic acid (ARA), remain debated. Almost all research to date has used fatty acid (FA) data expressed as percent of total FA (% total).

**Objective:** The objective of this study was to determine whether expressing fatty acid (FA) data as % of total or as absolute concentrations alters associations with clinical biomarkers.

**Methods:** Serum FA data obtained via electron capture negative-ion mass spectrometry was obtained from NHANES. Each FA was expressed both as % total and absolute concentration (µmol/L). Associations were examined between individual and total FAs and a panel of lipid and non-lipid biomarkers, including total cholesterol, LDL-C, HDL-C, triglycerides, blood pressure, body mass index, waist circumference, glucose, and insulin.

**Results:** Associations between LA and clinical biomarkers including triglycerides, cholesterol, HDL-C, BMI, glucose, and insulin, reversed direction depending on whether LA was expressed as % total or as a concentration. Similar reversals were observed for ARA, DHA, DPA, and stearic acid. Increases in total FA levels were accompanied by decreases in % total of several PUFAs and HUFAs, despite rising absolute concentrations. Total FA was positively associated, often strongly, with nine clinical markers and negatively associated with HDL-C.

**Conclusions:** Expression format significantly impacts observed FA associations. Reliance on % total FA values alone may misrepresent true associations between individual FAs and clinical endpoints, especially when the total fatty acid pool also changes size. To develop effective dietary guidance or clinical recommendations, it is essential to consider the underlying FA biology and total FA pool size when determining whether % total or absolute FA concentrations are more appropriate.

## Introduction

Fatty acids (FA) play essential roles in biological processes, and researchers have sought for decades to understand the relationships between dietary FA levels and health outcomes. Levels of saturated FAs, polyunsaturated fatty acids (PUFA), and highly unsaturated fatty acids (HUFA) have been associated with multiple disease phenotypes, including cardiovascular disease (CVD) and diabetes (1–4). Research into the importance and ideal quantities of individual FAs such as linoleic acid (LA), arachidonic acid (ARA), and eicosapentaenoic (EPA) remains active and the source of extensive debate (5–10).

Since the 1960s, FA quantitation data have primarily been generated by gas chromatography flame ionization detection (GC-FID), either in a percent of an individual FA (% total) relative to the sum of total FAs in all circulating lipid classes or in phospholipids. These data are typically presented as a % of the total FA pool being measured, and corresponding absolute FA concentrations have not typically been provided (1, 11–20). This is partially because peak-based chromatography data are naturally compositional (the areas under the individual peaks sum to the total area) and more precise, as obtaining concentrations requires the additional step of using standards to convert areas to concentrations.

This use of % total makes the data “compositional” as the proportions of total FA pool must sum to one. Due to this sum constraint, if one FA changes, the level of one or more other FAs must change typically in the opposite direction. Using absolute concentrations avoids this, as individual FA values can vary independently from one another but obscures the well understood competitive relationship between FA. However, it is often implicitly assumed that % total data can be treated like concentration data in that single FAs can be treated independently, and the overall association patterns would be the same as using concentrations. This is correct when the total pool of fatty acids is not changing, a condition which is not always satisfied. Sergeant et al. demonstrated that when comparing % total and concentrations, the associations between lipid based clinical biomarkers with PUFAs and HUFAs, such as LA and ARA, can change direction (21). While this group presented a mathematical framework determining the conditions for when associations reverse, the two data sets demonstrating these reversals empirically were limited in size and analyzed only lipid biomarkers.

This reversal is a well-characterized biological phenomenon. Gestational lipemia provides a clear illustration: the normal elevation of total circulating lipids relative to the non-pregnant state results in a decreased DHA percentage of total lipids, notwithstanding an increase in absolute DHA concentration (mg/dL plasma) (22).

The current study was prompted by the recognition that expressing FA data as concentrations versus % total can lead to differing conclusions, and by the observation that most published studies on circulating FAs and human health rely primarily on % total values (1, 11–20). The 2011–2012 National Health and Nutrition Examination Survey (NHANES) provided a unique opportunity to utilize mass spectrometry data to compare associations between circulating FAs expressed both as % total and as concentrations, across a large number of clinical endpoints in a nationally representative cohort. These analyses show that the direction and strength of associations between circulating FAs and health outcomes differ depending on the expression format, indicating a need to reassess dietary recommendations that have been based predominantly on % total FA data.

## Materials and Methods

### NHANES Data and Mass Spectrometry Analysis

Demographic, laboratory, and FA data from the 2011-2012 cycle were downloaded from the NHANES website. Following the data flowchart shown in **Supplementary Figure 1**, we analyzed data from 2,377 subjects (50% men, mean age 47 years). Briefly summarizing the NHANES protocol, participants were randomly selected among those in the fasting sub-sample (fasting at their morning examination) and provided a 0.5 mL blood sample. FAs were measured using the modified methods of Lagerstedt et al. at the Centers for Disease Control and Prevention or their collaborators/subcontractors’ laboratories (23, 24). Following alkaline hydrolysis, hexane extraction, and derivatization with pentafluorobenzyl bromide, FA esters were analyzed. Total FAs were hexane-extracted from the matrix (100uL serum or plasma), and an internal standard solution was applied to monitor FA recovery. Following the extract’s conversion to pentafluorobenzyl esters, the reaction mixture was injected into a capillary gas chromatograph column to separate individual FAs. Within 34 minutes, 30 individual FAs were detected with electron capture negative-ion mass spectrometry (Agilent GC model #7890A; MS model #5975C) and MSD Chemstation software (model: E.02.0049) by comparing peak analyte area of unknown to known in calibrator solution. Absolute concentrations (mmol/L) were determined by selected ion monitoring using ratios of stable-isotope-labeled deuterated internal standards. We then utilized these data to generate compositional data, % total for each FA.

### Statistical Modelling

All statistical analyses were conducted in R following the guidelines provided by the NHANES website (25, 26). The survey package was used to account for the NHANES sampling strategy (27). The distributions of outcome variables were inspected and log transformed where appropriate. Associations were determined either as Pearson’s correlation coefficient, or the regression coefficients from linear or logistic regression (svyglm) depending on the outcome type (continuous vs binary). Regression models were adjusted for age and sex.

## Results

### Associations of Linoleic Acid and Oleic Acid (Expressed as Percentage of Total Fatty Acids or Absolute Concentrations) with Common Clinical Biomarkers

Figure 1 shows the associations of LA, expressed as % total or absolute concentrations, with several lipid and non-lipid clinical biomarkers. The directions of the associations reverse depending on how LA is expressed. LA’s association with circulating triglycerides (TG) shows a marked reversal, while more subtle reversals are seen with HDL-C and total cholesterol (TC) (Figure 1A). Since LA is itself a lipid that primarily resides in complex lipids such as TG, cholesterol esters and phospholipids, it was also important to examine associations with non-lipid clinical biomarkers (Figure 1B). Associations between LA and systolic blood pressure, BMI, insulin and glucose levels all showed reversals when LA was expressed as a % total versus absolute concentrations. Specifically, the associations flipped from negative to positive when LA was expressed as absolute concentration.

**Figure 1.**
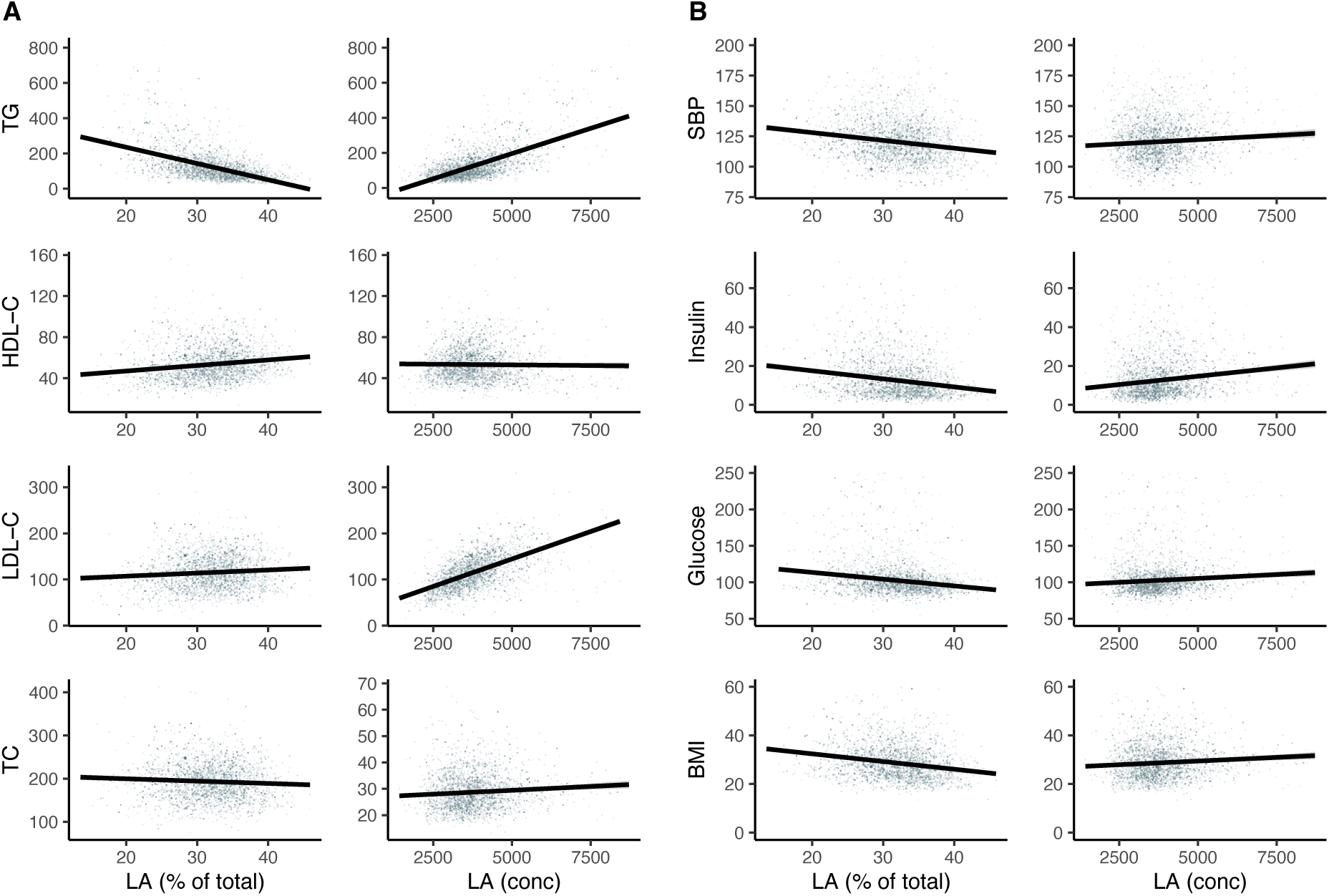
Four lipid-based (A) and four non-lipid (B) clinical biomarkers are expressed as a function of LA. LA is expressed as both % total and concentration (umol/L). Abbrev. TRG = triglycerides; Tot Chol = total cholesterol; SBP = systolic blood pressure; BMI = body mass index.

In contrast, for oleic acid (OA; 18:1, n-9), the associations were mostly consistent when the data were expressed as % total or absolute concentrations. **Supplementary Figure 1** shows the relationships between OA and the same clinical biomarkers examined in Figure 1. While the directions of the associations were more consistent with OA, there were still reversals in the direction with HDL-C and TC.

LA and OA were not the only FAs where the relationships reverse based on how they are expressed. **Table 1** shows which correlations reverse for 13 FAs and 9 clinical biomarkers when the data are expressed as % total versus absolute concentration. The most relationship reversals are observed with LA, ARA, docosapentaenoic acid (DPA; 22:5, n-6), DHA, and stearic acid (SA, 18:0) with nine, seven, nine, eight, and nine reversals, respectively. In many cases, the direction and magnitude of the correlation markedly change. Additionally, the significance of the FA variable in regression models may shift, such that the FA expressed as concentration was statistically significant, while its % total may not be, or vice versa.

**Table 1:**
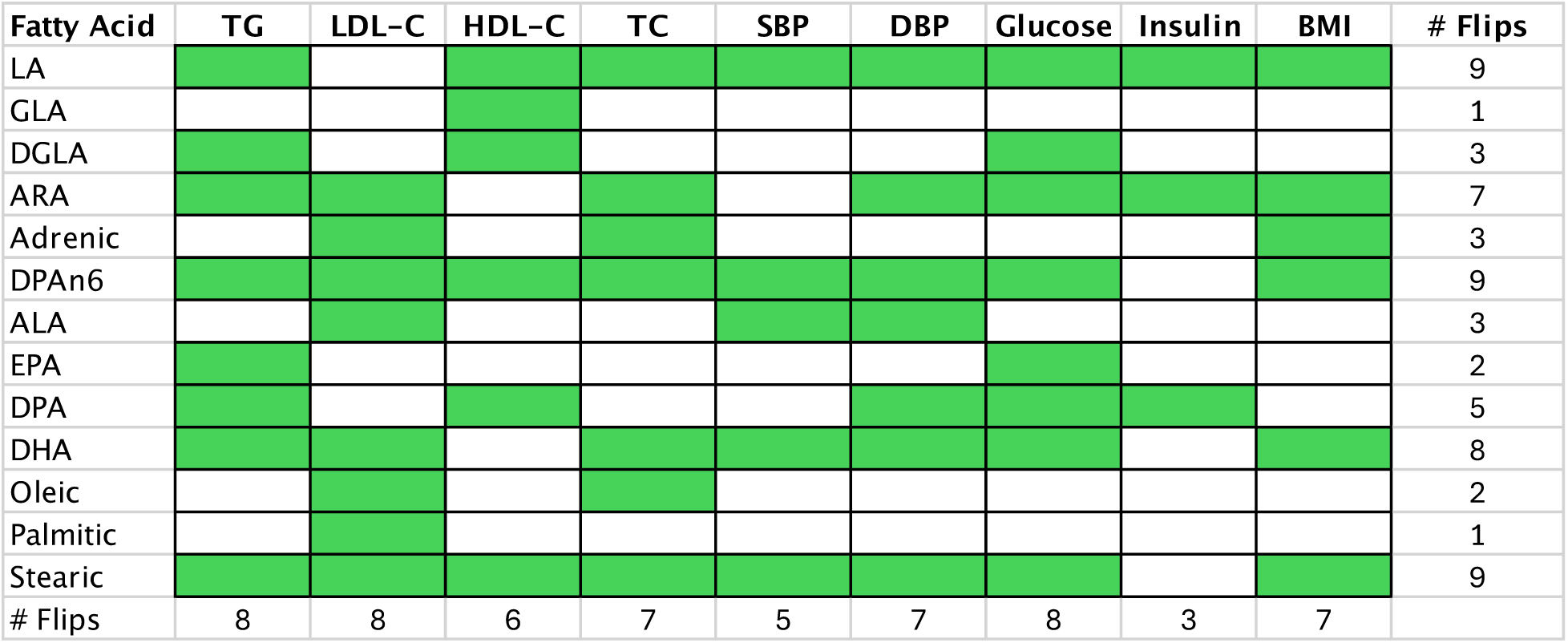
This table shows correlations between individual fatty acid (rows) and clinical biomarkers (columns). A green cell indicates that the association between % total and absolute concentrations reverse direction.

### Comparing % Total and Concentrations of Individual FAs in the Context of Total Circulating FAs

These data raised the important question of why do certain correlations reverse depending on whether they are expressed as % total or as FA concentrations? We hypothesized that like findings in studies of ARA and DHA in pregnant women, as the total concentration of circulating FAs increases, the relative percentage of some individual FAs such as ARA and DHA may decline if those FAs are not increasing as rapidly as others within the total FA pool. To test this hypothesis, we compared FA profiles, expressed both as % total and as concentrations, between individuals in the first (Q1) and fourth (Q4) quartiles of total circulating FA concentrations. The quantile-based approach using % total data has been widely applied in epidemiologic studies to examine associations between circulating FAs and clinical biomarkers or endpoints (1, 12–14, 16–18, 20).

Figure 2 illustrates that the overall average FA composition differs markedly between Q1 and Q4, whether expressed as % total or as absolute concentrations. For example, several PUFAs and HUFAs reverse direction depending on the reporting method used. For example, as total FA levels increase from Q1 to Q4, there are marked increases in the absolute concentrations of LA, ARA, OA, and palmitic acid. While increases in total FAs are reflected in the percentages of OA and PA, the changes are much smaller than the increases observed in their absolute concentrations. Importantly, although LA and ARA show a 1.6-fold and 1.7-fold increase in absolute concentrations, respectively, their % total values decrease by approximately 15%, highlighting how misleading % total data can be in the context of rising total FA levels.

**Figure 2.**
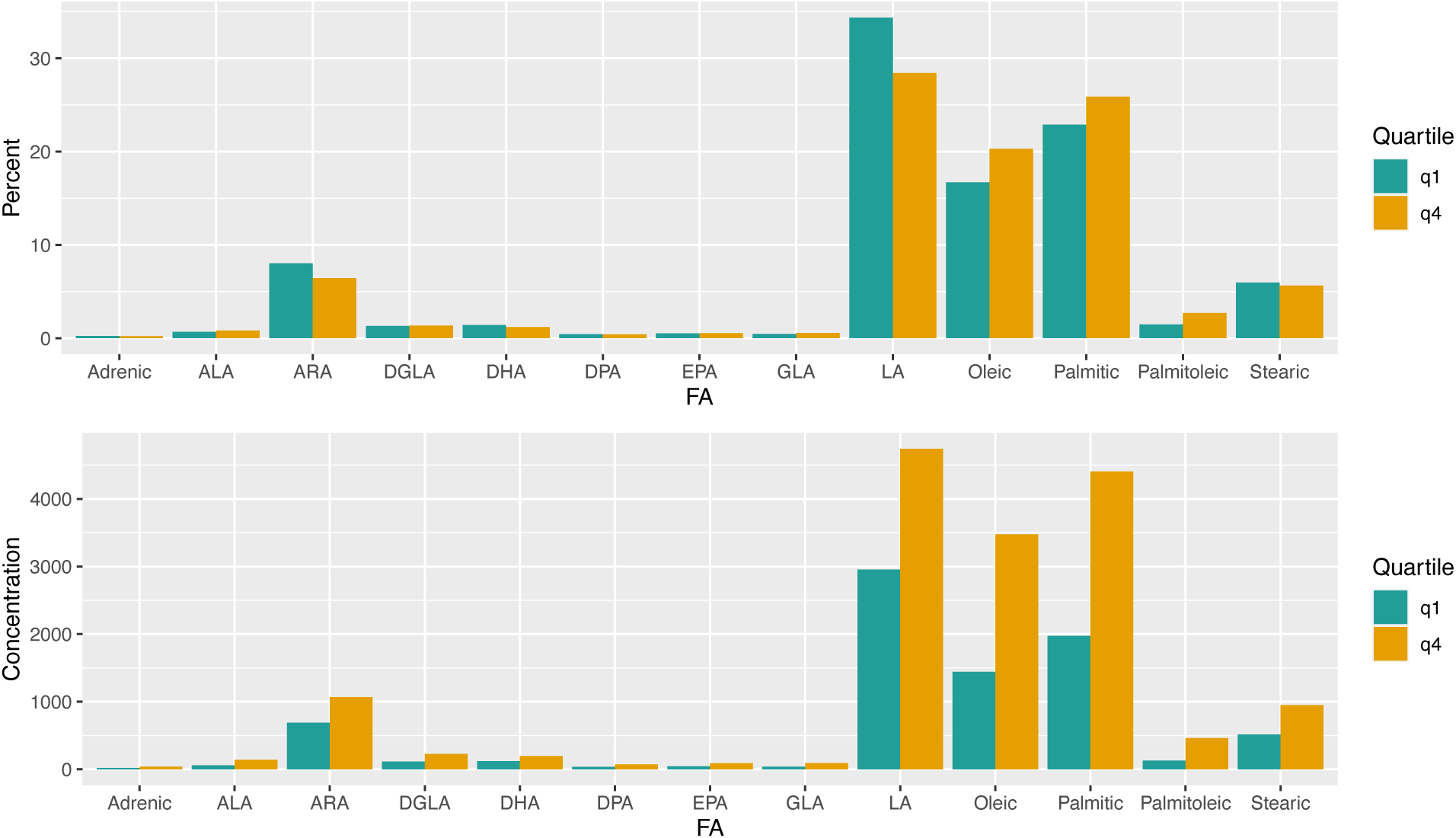
: FAs quantities expressed as % total (Top) or absolute concentrations (Bottom) in the first (q1) and fourth (q4) quartiles of total plasma FA levels.

Figure 3 further illustrates this phenomenon by showing relationships between total FA concentration and four specific FAs (LA, ARA, DHA, and EPA) expressed both as % total and as concentrations. For three of these (LA, ARA, and DHA), the direction of association with total FA reverses when switching from % total to concentration. That is, as total FA levels rise, these FAs decline in % total terms but increase in concentration. In contrast, EPA demonstrates a positive association with total FA levels in both measures, although the strength of association is considerably greater when using concentrations compared to % total.

**Figure 3.**
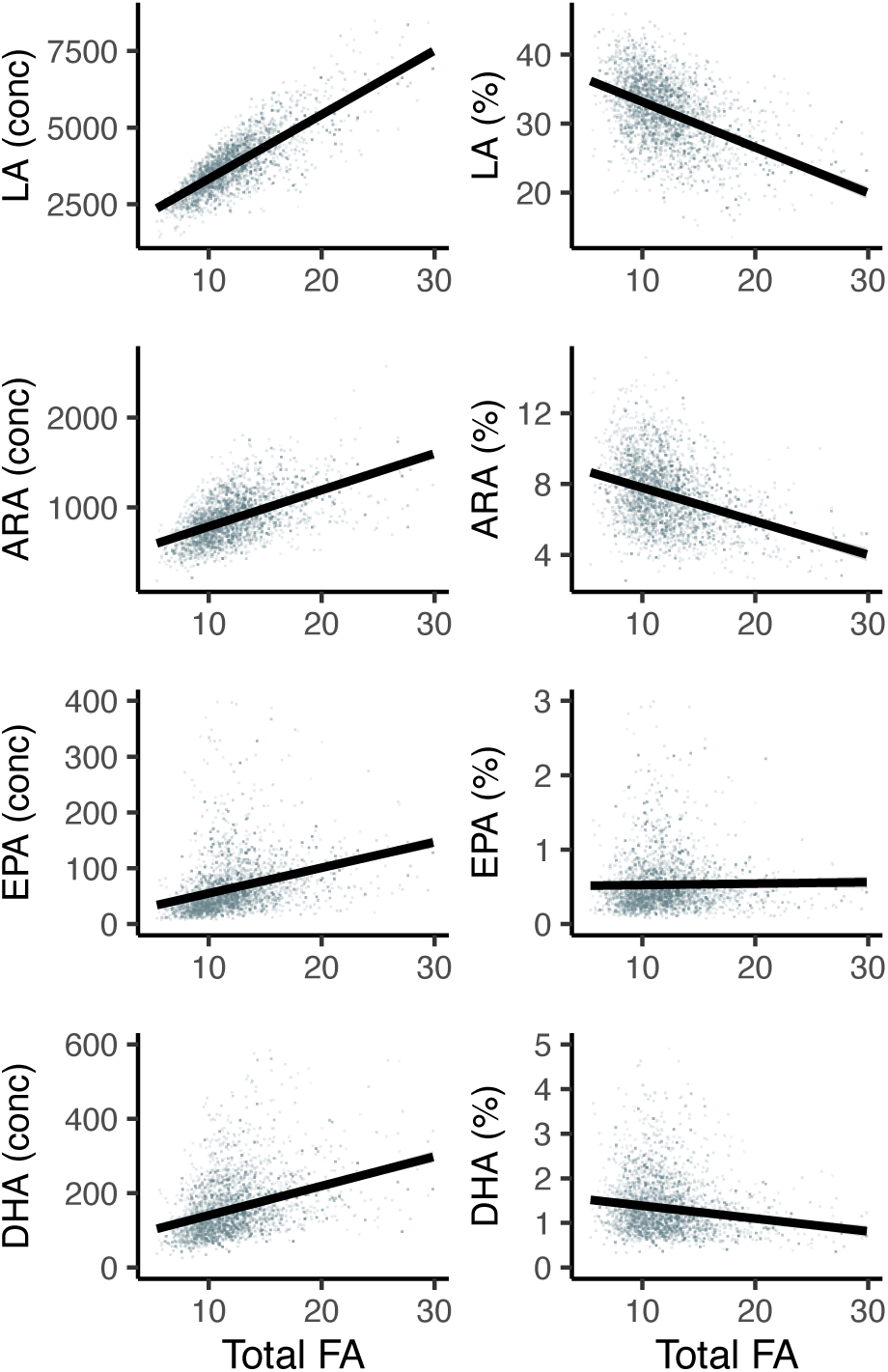
Plots of four individual FAs [expressed as both concentration (umol/L) and % total] plotted as a function of total FA concentration (mmol/L).

### Associations of Total Fatty Acids with Common Clinical Biomarkers

Figure 4 illustrates the significant associations between total FA and ten clinical biomarkers. Nine (TG, LDL-C, TC, insulin, glucose, systolic and diastolic blood pressure (BP), BMI, and waist circumference) were positively associated with total FA, while one, HDL-C was negatively associated with total FA. This highlights the importance of total FA and suggests that it should be considered in these types of analyses.

**Figure 4.**
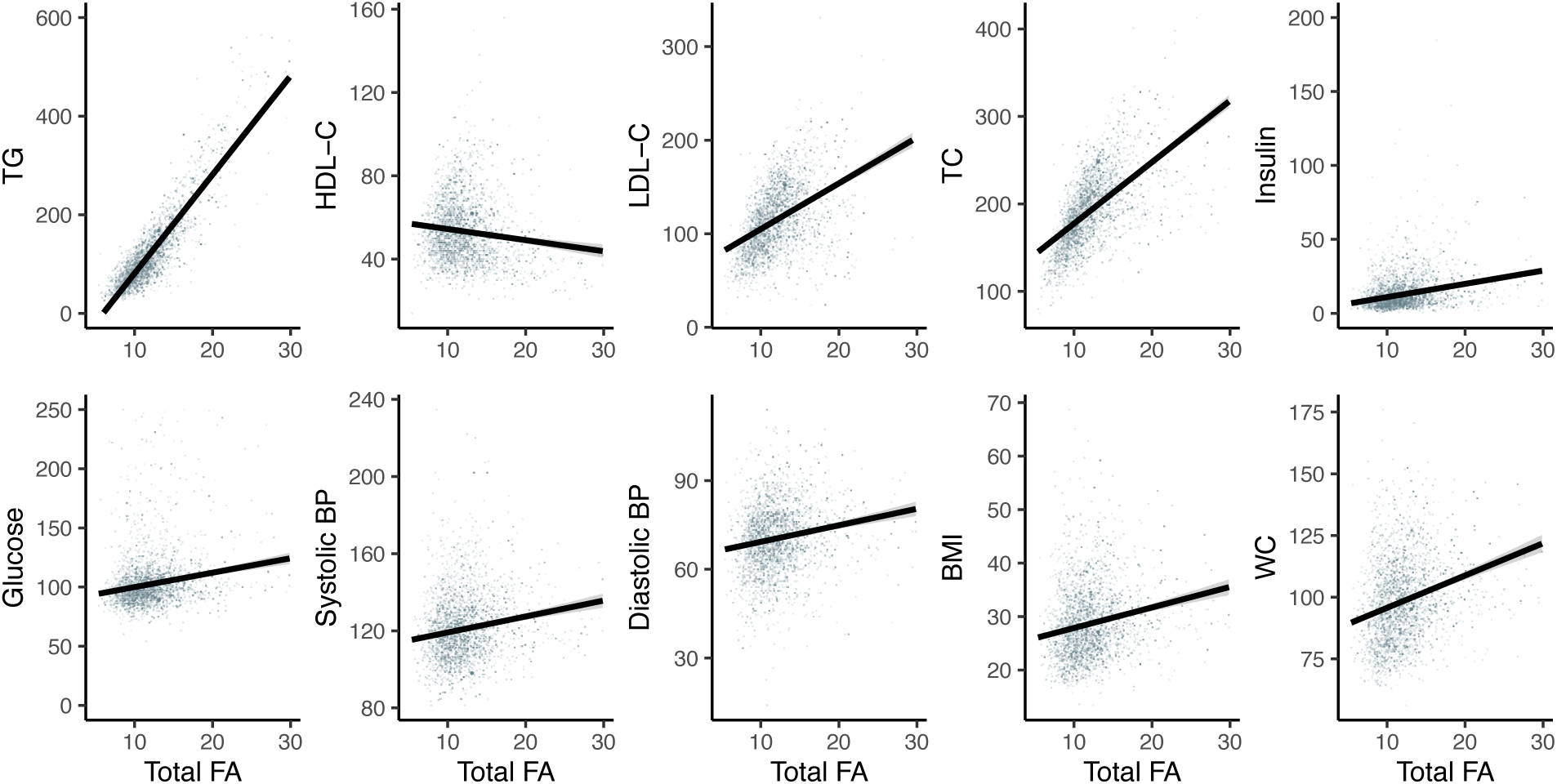
Clinical biomarkers plotted as a function of total FA concentrations. Abbreviations include: TRG, triglycerides; HDL, high-density cholesterol; LDL, low density cholesterol; TotChol, total cholesterol; BP, blood pressure; BMI, body mass index; waist circum, waist circumference.

## Discussion

An observed association between a potential cause and an outcome is more likely to reflect a true causal relationship when the link is strong, as stronger associations are less likely to be explained by confounding factors. The 2011–2012 NHANES, with its quantitative mass spectrometry data, extensive clinical biomarker panel, and large cohort, provided an important opportunity to compare associations between individual FAs and clinical biomarkers when FAs are expressed either as % total or as absolute concentrations. The findings of the current study reveal a critical issue in interpreting these associations and highlight the significance of expression format. Notably, we observed reversals in the direction of association for LA and several other FAs, including ARA, DPA, DHA, and SA, depending on whether values were expressed as % total or concentration. A particularly striking example is the reversal in the association between LA, the primary dietary n-6 PUFA, and nine lipid and non-lipid clinical biomarkers. These findings replicate an earlier study from our laboratory, which identified similar reversals in lipid-based biomarkers across two smaller cohorts (21).

With respect to clinical biomarkers, the current analysis calls into question both the strength and direction of previously reported associations and emphasizes the need for careful reconsideration of how circulating FAs are quantified and reported in epidemiological research (21, 28). These reversals also suggest that absolute concentrations of FAs can provide additional or alternative insights compared to using % total. In fact, total FA concentrations appear to be the best predictor of levels of clinical variables in many cases, but it is rarely considered, as total FA are not typically quantified.

While the current study focuses on clinical biomarkers, a primary question remains regarding the association between FAs and all-cause and coronary heart disease (CHD) mortality. Consistent with previous findings, a recent meta-analysis by Shi et al., which included 49 non-overlapping studies analyzing circulating FAs expressed as % total, reported significant inverse associations of n-3 HUFAs and LA with both CHD and stroke mortality (2). However, due to the limited availability of FA concentration data, it has only recently been possible to determine the relationship between absolute FA concentrations and all-cause, CHD, or stroke mortality. However, emerging evidence from large studies utilizing nuclear magnetic resonance (NMR) data to analyze FA concentrations suggests that the mode of FA expression may significantly influence observed associations. Notably, supplementary data (Table 13) from the Shi et al. study, which examined 139,538 individuals from the UK Biobank and INTERVAL cohorts, showed reversals in hazard (HR) ratios for CHD and stroke mortality depending on whether FAs were expressed as relative percentages or absolute concentrations (2). Additionally, a recent study by Zhang et al. (3) reported strong positive associations between the circulating n-6/n-3 PUFA and HUFA ratios and the risk of all-cause, cancer-related, and cardiovascular mortality.

The current study highlights a key limitation of observational research, namely, the challenge of drawing definitive conclusions about the relationships between individual FAs and clinical outcomes. While a randomized intervention trial with LA might seem warranted, such studies are often problematic due to the already high intake of dietary LA (∼17 g/day) in the standard Western diet. Substantially reducing this intake presents practical challenges related to the biological properties and availability of suitable replacement oils, as well as likely issues with participant compliance.

Conversely, the Sydney Diet Heart Study demonstrated that increased LA intake was associated with a marked rise in all-cause mortality and coronary deaths (29, 30). Given these complexities, both the feasibility and ethical justification for further randomized trials targeting LA remain uncertain, and thus, the existing RCT evidence regarding LA remains inconclusive.

Another important criterion is whether the observed associations are consistent with existing biological knowledge. LA and its metabolic product, ARA, are particularly relevant in this context given the capacity of ARA to be converted to pro-inflammatory and pro-thrombotic oxylipins (31, 32). Additionally, high LA intake is known to compete with alpha-linolenic acid (ALA, an n-3 PUFA), thereby reducing the biosynthesis of n-3 HUFAs such as EPA and DPA and their oxylipin derivatives (33–35). This reduction may in turn lead to the reduction of anti-inflammatory, anti-thrombotic, pro-resolving oxylipins. Taken together, these mechanistic insights support the plausibility that excessive dietary LA could contribute to the pathogenesis of chronic inflammatory conditions and cardiovascular disease. The balance between n-6 and n-3 PUFA-derived oxylipins may therefore be a critical factor in maintaining health, and disruptions to this balance, driven by disproportionately high LA intake, may help explain the associations observed in both biomarker and mortality studies.

These findings have significant implications for both research and clinical practice. The inappropriate use of percentage-based FA data, while common, may have inadvertently obscured critical associations and led to incomplete and more importantly, misleading conclusions. For example, recommendations to limit DHA intake during pregnancy based on its apparent decline when expressed as % total would have been clearly misguided. Similarly, it is worth questioning whether the rationale for increasing dietary LA, based on % total circulating LA being associated with lower cardiometabolic risk, disease, or mortality, may have also been flawed. Future studies should incorporate both expression methods and consider total pool size to provide a more comprehensive understanding of FA-related health outcomes (36). This dual-approach framework will improve the accuracy of dietary recommendations and deepen our understanding of FA biology in both health and disease.

## Data Availability

All data used are available online at https://wwwn.cdc.gov/nchs/nhanes/continuousnhanes/default.aspx?BeginYear=2011

## Acknowledgements

**Brian Hallmark**: Conceptualization, Methodology, Investigation, Software, Formal Analysis, Writing - Original Draft <u>and</u> Review & Editing, Visualization. **Manja Zec**: Conceptualization, Methodology, Investigation, Writing - Original Draft <u>and</u> Review & Editing **Laurel Johnstone**: Project administration, Resources **Carrie S. Standage-Beier**: Data Curation, Resources **Susan Sergeant**: Writing - Review & Editing **Justin Snider**: Investigation, Writing - Review & Editing **J. Thomas Brenna**: Writing - Review & Editing **Floyd H. Chilton**: Conceptualization, Writing - Original Draft <u>and</u> Review & Editing, Supervision, Funding acquisition

---

**Supplementary Figure 1.**
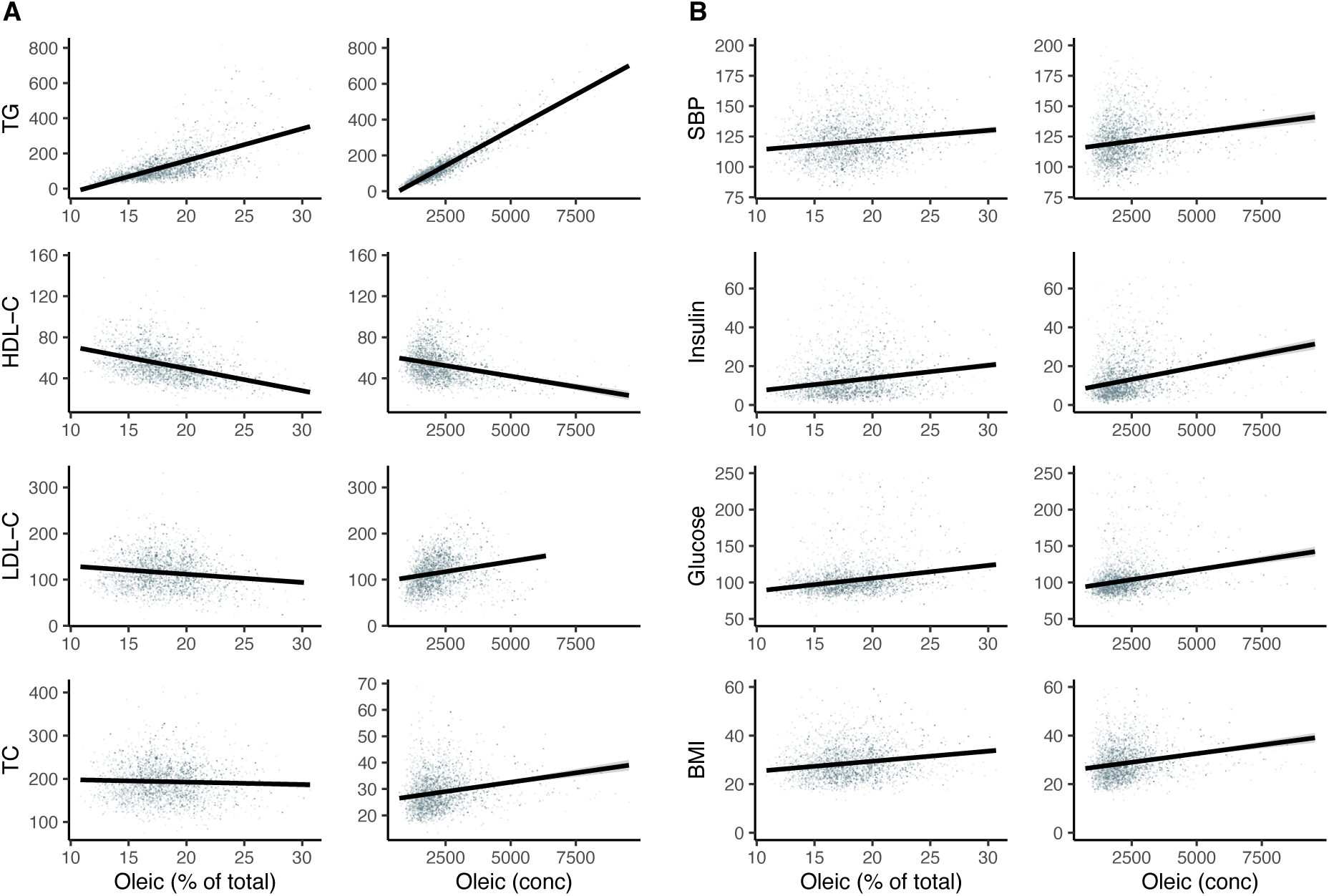
(A) Four lipid-based and (B) four non-lipid clinical biomarkers are expressed as a function of oleic acid (OA). OA is expressed as both % total and concentration (umol/L). Abbreviations include: TRG, triglycerides; HDL, high-density cholesterol; LDL, low density cholesterol; TotChol, total cholesterol; BP, blood pressure; BMI, body mass index; waist circum, waist circumference.

**Supplementary Figure 1.**
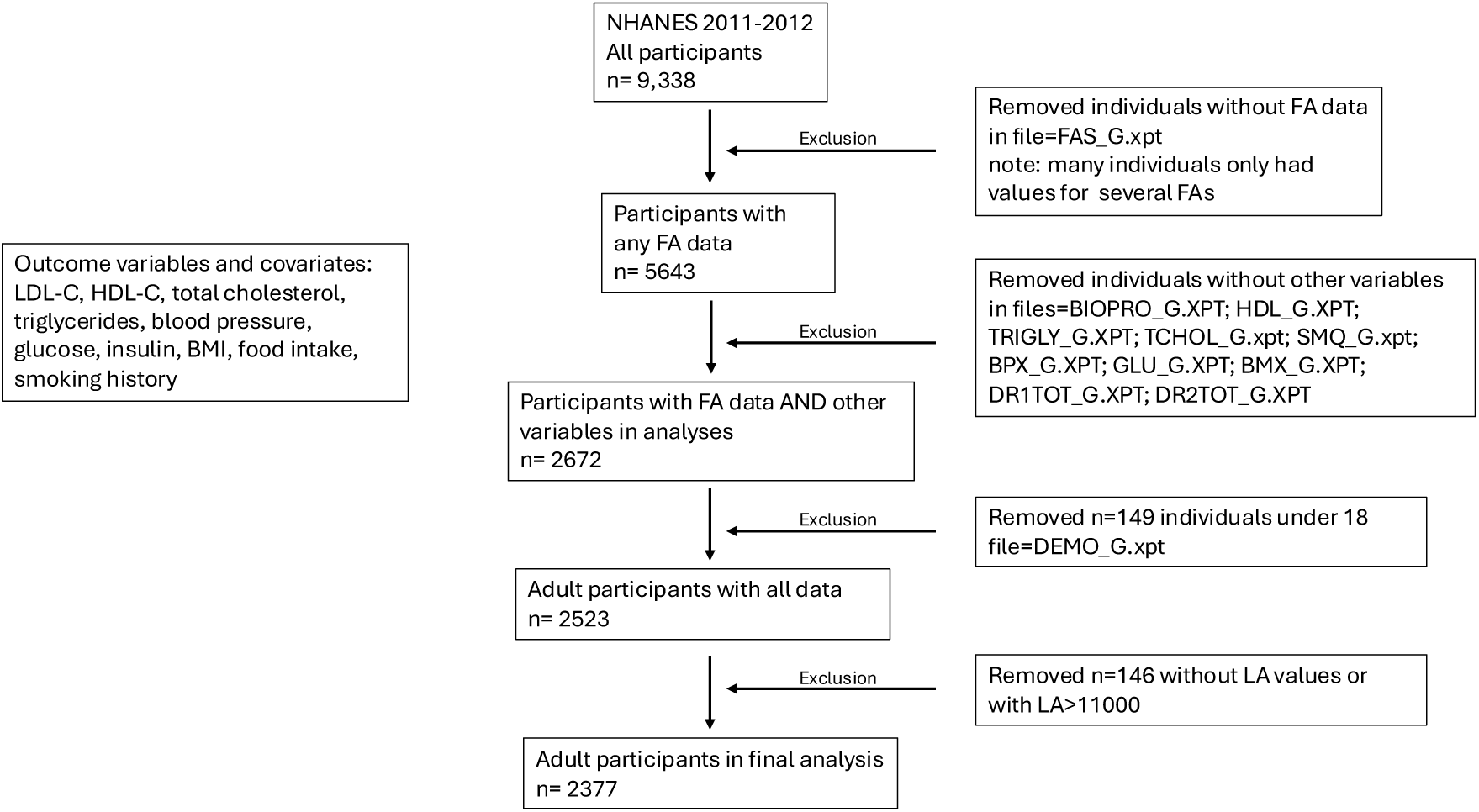
Flowchart showing data selection from NHANES 2011-2012.

